# The risk of household secondary invasive Group A Streptococcal infections after a prophylaxis policy change in the Netherlands

**DOI:** 10.1101/2025.07.07.25330994

**Authors:** Brechje de Gier, Bart J.M. Vlaminckx, Annika van Roon, Mirjam J. Knol, Margreet J.M. te Wierik, Daan W. Notermans, Hester E. de Melker, Nina M. van Sorge

## Abstract

**Importance:** Household contacts of patients with invasive group A streptococcal (iGAS) disease have an increased risk of iGAS. In the Netherlands, the iGAS public health policy was changed in January 2023, offering antibiotic prophylaxis to household contacts of all iGAS patients rather than only those presenting with necrotising fasciitis or streptococcal toxic shock syndrome.

**Objective:** To estimate risk of iGAS in the general population and among household- and other contacts of primary iGAS patients, before and after the policy change.

**Design:** A nationwide open cohort, linking population registry data with iGAS laboratory data, for the study period April 2022-December 2024.

**Setting:** Population-based.

**Participants:** All persons included in the Dutch population registry at any time during the study period. The case definition was an iGAS isolate submitted to the Netherlands Reference Laboratory for Bacterial Meningitis, with disease onset in the study period.

**Exposure:** For contacts of primary iGAS patients, exposure risk period was defined as the 30 days after culture date of the index patient. Exposure under the new policy was defined as all person-time after 20 January 2023.

**Main Outcomes and Measures:** We estimated the incidence rate ratio (IRR) of iGAS during the 30-day risk period compared to unexposed person-time. Secondary attack rates among household contacts were estimated, calculating an odds ratio (OR) to compare attack rates before and after the policy change. Estimates were adjusted for age group, sex, household socioeconomic status and year-quarter.

**Results:** A total of 19,006,247 persons contributed 51,067,977 person-years to the analysis. A total of 3,644 iGAS isolates from 3,630 unique persons was included, of which 14 were household secondary cases. The IRR for household contacts during the risk period was 235.25 (95%CI 94.35-586.59) before, and 74.00 (95%CI 35.17-155.71) after the policy change, compared to unexposed person-time. Secondary attack rate among household contacts was 0.219% (n=7) before, and 0.047% (n=7) after the policy change, adjusted OR 0.17 (95%CI 0.03-0.83).

**Conclusions and relevance:** In this nationwide cohort study, we observed a reduction in secondary iGAS risk among household contacts after implementation of an expanded antibiotic prophylaxis policy.

## Introduction

*Streptococcus pyogenes* or Group A streptococcus (GAS) is a human-specific Gram-positive bacterium, that can be carried asymptomatically but also causes a large burden of non-invasive and invasive disease. Known risk factors for invasive group A streptococcus (iGAS) infections include young (0-5 years) or old (60+ years) age, having underlying medical conditions, the peripartum period, and co-infections such as influenza or chickenpox [1]. Although iGAS is uncommon, a surge of iGAS has been observed globally after lifting all societal restrictions related to the COVID-19 pandemic. iGAS has a high case fatality, estimated between 8 and 16% [1]. Close contacts of an iGAS patient have a greatly increased risk of developing iGAS, with estimates ranging between 800 to over 5000 per 100,000 person-years at risk, compared to background incidences of 2-4 per 100,000 person-years [1-3]. In the absence of effective vaccines, the only available public health measure to prevent iGAS is antibiotic prophylaxis for individuals after potential high-risk GAS exposure. There is currently no clear consensus about how to most effectively prevent secondary iGAS infection.

In 2008, three clinical presentations of iGAS became notifiable by law in the Netherlands: streptococcal toxic shock syndrome (STSS), necrotising fasciitis and puerperal sepsis. Household contacts of patients with STSS or necrotising fasciitis were offered antibiotic prophylaxis to prevent secondary iGAS infection. In the Netherlands, a more than twofold increase in the annual number of notifiable iGAS infections was observed in 2022 compared to the annual average in the pre-COVID-19 pandemic years, leading to an estimated incidence of 10 per 100,000 per year [4, 5]. We have observed an absolute and relative increase of the globally disseminated *emm*1 clade M1_UK_, as well as the emergence of new variants of concern such as *emm*1.134 in 2022-2023, followed by an expansion of a previously rare *emm* type 3.93 [5, 6]. A case-only analysis of the Dutch data showed that these three *emm* types more often clustered within households, suggesting an increased secondary attack rate [unpublished data]. These observations prompted a change in the Dutch public health policy regarding antibiotic prophylaxis on 19 January 2023 where household contacts of all iGAS patients were eligible for antibiotic prophylaxis to prevent secondary iGAS infection [7]. To implement this policy, all iGAS infections (i.e. all iGAS compatible disease presentations with *S. pyogenes* detected in a normally sterile site) became notifiable by law. It was decided that a single daily dose of 500 mg azithromycin for three days was the preferred prophylaxis regimen to optimize acceptability and adherence. However, as macrolide resistance is not rare among *S. pyogenes* isolates [4], the prophylaxis can be changed based on the antibiotic susceptibility profile of the primary case. Alternative prophylactic regimes include either a 10-day course of clindamycin or oral penicillin in combination with four days of rifampicin. Both household- and other close contacts are informed about their increased risk and are advised to seek medical care without delay in case of any symptoms consistent with (i)GAS infection, to enable early treatment.

The main aim of this retrospective population-based cohort study was to assess the impact of the policy change on the risk of secondary iGAS infection among household- and other close contacts. We further describe the population incidence and risk factors for iGAS, and 30-day mortality after iGAS, in the Netherlands in the post-pandemic years 2022-2024.

## Methods

### Data sources

The Netherlands Reference Laboratory for Bacterial Meningitis (NRLBM) has collected *S. pyogenes* isolates for nationwide bacteriological surveillance of iGAS since April 2022. All medical microbiological laboratories in the Netherlands are requested to submit iGAS isolates to the NRLBM, when cultured from a normally sterile site or from a non-sterile site in combination with a clinical presentation of iGAS. While submission is voluntary, the number of isolates received from unique patients during a time period when all iGAS was notifiable exceeded the number of iGAS notifications (2,834 isolates versus 2,492 notifications between February 2023 and December 2024). Also, the number of GAS isolates cultured from blood submitted to the NRLBM slightly exceeded the number of GAS blood cultures in the national antimicrobial susceptibility surveillance system ISIS-AR in 2023 (Van der Putten et al, unpublished results). All iGAS isolates are *emm* (sub)typed according to the CDC protocol through PCR amplification and subsequent Sanger sequencing of the 180-bp domain that contains the 50 hypervariable codons [5]. Submitted isolates are accompanied by limited information regarding patient characteristics, including sex, birth date and postal code. Through the Statistics Netherlands (CBS) microdata system, isolates were linked to population registry data. Data on birth dates and sex were retrieved from the population registry.

## Definitions

An iGAS case was defined as a person with a linked iGAS isolate during the study period. A composite score of socioeconomic status, based on income, wealth and occupation status, was available at the household level. Household compositions, with start and end dates, were used to identify household contacts and socioeconomic status per participant. These households can be private or institutional (e.g. long term care facilities for disabled or elderly care). Household contacts were defined as a person registered as belonging to the same household as a primary case at the iGAS disease onset date (the first known date of culture specimen or isolate receipt by the NRLBM). For clusters of co-primary cases, i.e. contacts with the same index date, the primary and secondary case labels were assigned at random and one of the two was included as outcome event in the main analysis. For children born since 2022 and therefore not included in the person-network datasets, we assumed the non-household relatives of their household contacts were also the non-household relatives of the children. Non-household relatives, colleagues, classmates and neighbors were identified through the person-network datasets provided by Statistics Netherlands [8-11]. As these datasets were available for the period up to and including 2022, we assumed the contacts in 2022 were still their contacts in 2024. The period before the policy change was defined as 1 April 2022-19 January 2023, and compared to the period 20 January 2023-31 December 2024, when the extended antibiotic prophylaxis policy was in effect.

### Ethics

The RIVM Centre for Clinical Expertise verified whether this study complies with the Dutch law for Medical Research Involving Human Subjects (WMO) or with the EU Clinical Trial Directive (2001/20/EC), and was of the opinion that review by an ethical research committee or institutional review board is not necessary by current national and European legislation (study number EPI-692).

### Analytical methods

An open cohort approach was used, where persons were included from 1 April 2022 or their birth date (whichever came last) and followed until 31 December 2024, their first iGAS episode, or death, whichever came first. When multiple isolates were received from the same person within 30 days, these were considered as belonging to the same episode. As primary outcome, the first iGAS episode per person was used. Persons with multiple iGAS episodes could however contribute to multiple exposure periods for their contacts. The primary exposure of interest was being a contact of an iGAS patient, during the 30-day risk period after the primary patient’s index date, under either the old or the new prophylaxis policy. The risk period was truncated in case of an event, therefore only the days leading up to the event of the contact are included in the person-time denominators. For secondary attack rate estimates, the number of contacts was the denominator.

Negative binomial regression was used to estimate IRR of contacts during the risk period compared to the population background incidence. IRRs were compared before and after the policy change, and an interaction term between period and the exposure was used to test for significance. Logistic regression was used to compare secondary attack rates (SAR) among household contacts before and after the guideline change, and to estimate risk of 30-day mortality. All models were adjusted for age group, sex, calendar time (in year-quarters), and household socioeconomic status (SES) in quintiles (where quintile 1 represents the lowest SES). In the model comparing SAR among household contacts, standard errors were adjusted for clustering by household. Analyses were performed in R, using packages tidyverse, Epi, mgcv, lmtest and multiwayvcov.

Results are based on calculations by the RIVM, in project number 9892, using non-public microdata from Statistics Netherlands. As per the Statistics Netherlands policy, no numbers below 5 or estimates based on fewer than 5 events could be reported.

The STROBE reporting guideline for cohort studies was used.

## Results

Figure 1 shows the number of S. *pyogenes* isolates from iGAS patients submitted to the NRLBM during the study period by *emm* type. During the 2022-2023 season, *emm*1.0 dominated, while *emm*3.93 clearly dominated during the 2023-2024 season, as described previously [5, 6]. A total of 4,215 iGAS isolate records were submitted to Statistics Netherlands, of which 3,729 (88.5%) could be deterministically linked to unique persons in the population registry (Supplementary Figure 1). Linkage was significantly more often possible for ages 46 years and older, females, and isolates of *emm*4.0, but did not differ by policy period (Supplementary Table 1). Eighteen isolates were excluded since they were collected before the start of the study period, and a further 67 isolates were excluded when de-duplicating isolates from the same person within 30 days. After exclusions, 3,644 iGAS isolates from 3,630 unique persons were included in the analysis.

**Figure 1.**
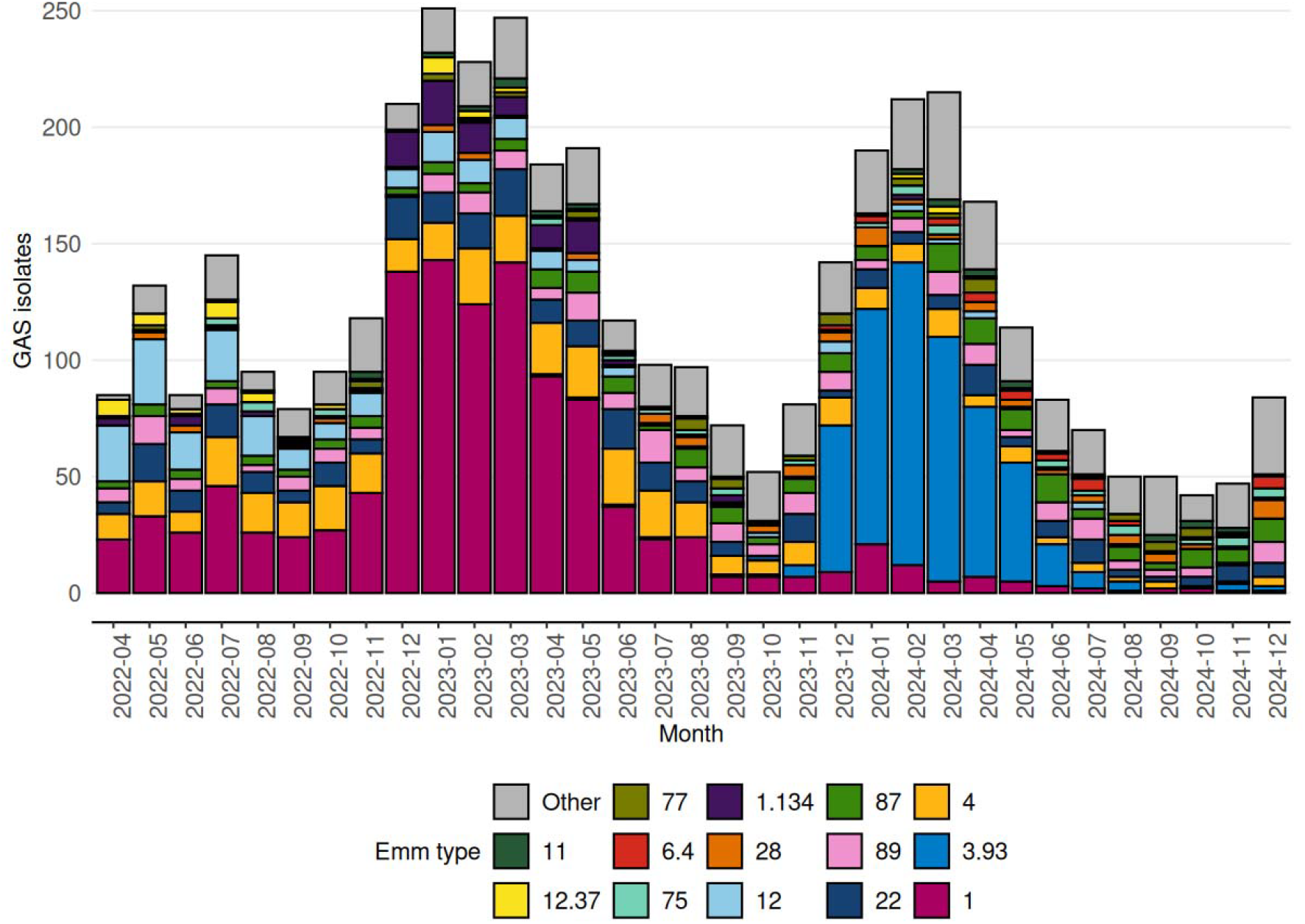
Number of S. *pyogenes* isolates received and typed by the Netherlands Reference Laboratory for Bacterial Meningitis during the study period 1 April 2022-31 December 2024, by month and *emm* type (n=4,129).

A total of 19,006,247 persons contributed 51,067,977 person-years at any time during the study period. Of this population, 49.8% was registered as male (Supplementary Table 2). Over the complete study period, 3,609/3,630 (99.4%) were primary iGAS cases, i.e. cases not occurring during the risk period of a known contact (Table 1), corresponding with a population background incidence of 7.07 per 100,000 person-years. The incidence peaked in the first quarter of 2023 at 14.15 per 100,000 person-years (Table 1). Age groups 0-5 and 66+ years had a significantly higher incidence compared to age group 45-65 years. Persons registered as male had a significantly lower incidence compared to persons of female or unknown sex. This is likely related to the risk of puerperal fever or -sepsis for women of childbearing age (see Supplementary Figure 1). Only the fourth quintile (second-to highest) of household SES had a significantly higher incidence compared to the first quintile. Incidences by year-quarter reflect the winter seasonality of iGAS.

**Table 1.** Number of person-years (rounded to integer), iGAS events, incidence of iGAS per 100,000 person-years and by exposures of interest, 1 April 2022-31 December 2024, the Netherlands. Incidence rate ratios (IRR) with 95% confidence interval (CI), adjusted for age group, sex, household socioeconomic quintile and year-quarter.

|  |  | Person-years | iGAS events | Incidence per 100,000 person-years | IRR (95% CI) |
| --- | --- | --- | --- | --- | --- |
| **Exposure** | None | 51,033,734 | 3,609 | 7.07 | [ref] |
|  | Household | 1,451 | 14 | 964.79 | **113.14 (65.49-195.46)** |
|  | Non-household relatives | 7,852 | <5 |  |  |
|  | Colleagues and classmates | 11,178 | <5 |  |  |
|  | Neighbors | 13,762 | <5 |  |  |
| **Age group (years)** | 0-5 | 3,011,755 | 355 | 11.79 | 1.98 (1.71-2.29) |
|  | 6-19 | 7,733,534 | 225 | 2.91 | 0.50 (0.42-0.59) |
|  | 20-45 | 17,521,805 | 1,128 | 6.44 | 1.05 (0.94-1.17) |
|  | 46-65 | 13,381,750 | 790 | 5.90 | [ref] |
|  | 66+ | 9,419,133 | 1,132 | 12.02 | 2.13 (1.89-2.41) |
| **Sex** | Female or unknown | 25,635,174 | 1,952 | 7.61 | [ref] |
|  | Male | 25,432,804 | 1,678 | 6.60 | 0.86 (0.79-0.94) |
| **Household socioeconomic status quintile** | 1 (lowest) | 9,889,087 | 735 | 7.43 | [ref] |
|  | 2 | 9,921,409 | 778 | 7.84 | 1.00 (0.88-1.13) |
|  | 3 | 9,953,440 | 713 | 7.16 | 1.08 (0.95-1.22) |
|  | 4 | 9,969,159 | 689 | 6.91 | 1.14 (1.00-1.30) |
|  | 5 | 9,973,539 | 670 | 6.72 | 1.13 (0.99-1.29) |
|  | unknown | 1,361,345 | 45 | 3.31 |  |
| **Year-quarter** | 2022-Q2 | 4,670,196 | 278 | 5.95 | [ref] |
|  | 2022-Q3 | 4,671,408 | 279 | 5.97 | 0.98 (0.80-1.2) |
|  | 2022-Q4 | 5,635,926 | 524 | 9.30 | 1.48 (1.24-1.78) |
|  | 2023-Q1 | 3,605,008 | 510 | 14.15 | 2.16 (1.80-2.59) |
|  | 2023-Q2 | 4,618,864 | 440 | 9.53 | 1.44 (1.20-1.74) |
|  | 2023-Q3 | 4,670,868 | 236 | 5.05 | 0.82 (0.66-1.01) |
|  | 2023-Q4 | 4,671,280 | 248 | 5.31 | 0.86 (0.70-1.06) |
|  | 2024-Q1 | 4,618,692 | 556 | 12.04 | 1.84 (1.53-2.20) |
|  | 2024-Q2 | 4,617,811 | 316 | 6.84 | 1.03 (0.85-1.26) |
|  | 2024-Q3 | 4,669,237 | 153 | 3.28 | 0.52 (0.41-0.66) |
|  | 2024-Q4 | 4,618,688 | 90 | 1.95 | 0.32 (0.24-0.41) |

In total, 14 secondary iGAS cases were identified among household contacts of primary iGAS patients during the 30-day risk period, corresponding to an incidence of 964.79 per 100,000 person-years. The adjusted incidence rate ratio (IRR) for household contacts in the 30-day risk period compared to the population incidence was 113.14 (95%CI 65.49-195.46) over the entire study period. For non-household relatives, colleagues and classmates, and neighbors, the number of secondary cases was too low (<5) to present estimates according to the Statistics Netherlands policy.

Overall 30-day mortality was 10.2%, and significantly lower for cases with *emm* type 4.0 compared to *emm* type 1.0 (4.7% versus 12.4%, OR 0.47, 95%CI 0.26-0.78, Table 2). Mortality was significantly lower for ages 0-45 years compared to 46-65 years, and significantly higher for 66+. No significant difference in mortality was observed between sexes, socioeconomic quintiles or year-quarters (Table 2).

**Table 2.** 30-day mortality of iGAS cases between 1 April 2022-31 December 2024, the Netherlands, by emm type and odds ratios (OR) with 95% confidence interval (CI), adjusted for agegroup, sex, household socioeconomic quintile and year-quarter.

|  |  | iGAS cases | Died within 30 days, n (%) | OR (95% CI) |
| --- | --- | --- | --- | --- |
|  | Overall | 3,630 | 369 (10.2) |  |
| **emm type** | 1.0 | 1,030 | 128 (12.4) | [ref] |
|  | 12.0 | 192 | 25 (13.0) | 0.99 (0.58-1.65) |
|  | 22.0 | 266 | 21 (7.9) | 0.73 (0.42-1.22) |
|  | 3.93 | 523 | 72 (13.8) | 1.04 (0.65-1.65) |
|  | 4.0 | 362 | 17 (4.7) | 0.47 (0.26-0.78) |
|  | 87.0 | 173 | 12 (6.9) | 0.62 (0.31-1.17) |
|  | 89.0 | 187 | 18 (9.6) | 0.75 (0.40-1.32) |
|  | Other emm types | 897 | 76 (8.5) | 0.66 (0.46-0.93) |
| **Age group** | 0-5 | 355 | 16 (4.5) | 0.49 (0.27-0.84) |
|  | 6-19 | 226 | 7 (3.1) | 0.33 (0.14-0.68) |
|  | 20-45 | 1,127 | 27 (2.4) | 0.29 (0.18-0.45) |
|  | 46-65 | 792 | 71 (9.0) | [ref] |
|  | 66+ | 1,130 | 248 (21.9) | 2.73 (2.04-3.70) |
| **Sex** | Female or unknown | 1,952 | 165 (8.5) | [ref] |
|  | Male | 1,678 | 204 (12.2) | 1.16 (0.92-1.46) |
| **Household socioeconomic status** | 1 (lowest) | 735 | 94 (12.8) | [ref] |
|  | 2 | 778 | 116 (14.9) | 1.10 (0.81-1.50) |
|  | 3 | 713 | 73 (10.2) | 1.05 (0.75-1.49) |
|  | 4 | 689 | 48 (7.0) | 0.95 (0.64-1.39) |
|  | 5 | 670 | 32 (4.8) | 0.83 (0.52-1.29) |
|  | unknown | 45 | 6 (13.3) |  |
| **Year-quarter** | 2022-Q2 | 278 | 20 (7.2) | [ref] |
|  | 2022-Q3 | 279 | 30 (10.8) | 1.58 (0.85-2.99) |
|  | 2022-Q4 | 524 | 66 (12.6) | 1.51 (0.87-2.72) |
|  | 2023-Q1 | 510 | 51 (10.0) | 1.16 (0.66-2.13) |
|  | 2023-Q2 | 440 | 36 (8.2) | 0.96 (0.53-1.80) |
|  | 2023-Q3 | 236 | 17 (7.2) | 1.15 (0.56-2.35) |
|  | 2023-Q4 | 248 | 23 (9.3) | 1.23 (0.62-2.44) |
|  | 2024-Q1 | 556 | 70 (12.6) | 1.43 (0.78-2.71) |
|  | 2024-Q2 | 316 | 36 (11.4) | 1.21 (0.64-2.36) |
|  | 2024-Q3 | 153 | 13 (8.5) | 1.11 (0.50-2.42) |
|  | 2024-Q4 | 90 | 7 (7.8) | 1.02 (0.37-2.55) |

### Impact of policy change

In the period before the policy change (1 April 2022-19 January 2023), 7 secondary cases were identified among 3,195 household contacts, resulting in an IRR of 235.25 compared to the population incidence (95%CI 94.35-586.59; Table 3). After the policy change on 20 January 2023, there was a significant reduction in the IRR to 74.00 (95%CI 35.17-155.71; p for interaction 0.016), also based on 7 secondary cases among 14,974 household contacts. The population background incidence and the median number of household contacts per primary case did not differ between the two periods. Secondary attack rates (SAR) among household contacts were 0.219% before, and 0.047% after the guideline change (aOR 0.17, 95%CI 0.03-0.83).

**Table 3.** iGAS infections in households in the Netherlands, stratified by periods with the old and the new policy in place (1 April 2022-19 January 2023 and 20 January 2023-31 December 2024, respectively) . [1] Incidence rate ratio (IRR) adjusted for age group, gender, household socioeconomic quintile and year-quarter.

|  | **Old policy (1 April 2022- 19 January 2023)** | **New policy (20 January 2023- 31 December 2024)** |
| --- | --- | --- |
| **Primary iGAS cases in multi-person households** | 927 | 2,213 |
| **Household contacts** | 3,195 | 14,974 |
| **Median household contacts per case (IQR)** | 2 (1-3) | 2 (1-3) |
| **Secondary iGAS cases among household contacts** | 7 | 7 |
| **Secondary attack rate** | 0.219% | 0.047% |
| **Person-years household contacts (rounded to integer)** | 234 | 1217 |
| **Incidence among household contacts per 100,000 person-years** | 2995 | 575 |
| **Background population incidence per 100,000 person-years** | 7.17 | 7.03 |
| **IRR compared to unexposed <sup>[1]</sup>** | 235.25 (95% CI 94.35-586.59) | 74.00 (95% CI 35.17-155.71) |

## Discussion

We observed a significant reduction in risk of secondary iGAS infection for household contacts after a policy change, which extended antibiotic prophylaxis eligibility to household contacts of all iGAS patients rather than only household contacts of primary cases with STSS or necrotizing fasciitis. This reduction was visible both as a 4.5-fold reduction in SAR, as well as a three-fold difference in IRR, where the person-time denominator was truncated in case of an event. Data was not available on the actual prescription or uptake of antibiotic prophylaxis, therefore we could not estimate direct effectiveness of the prophylaxis against secondary iGAS infection.

Based on a 2023 study in the Netherlands, we know that there was a high overall acceptance of antibiotic prophylaxis with an uptake of 95% among eligible contacts. Of these contacts, 4.4% developed symptoms consistent with mild GAS infection, but no secondary iGAS infection was observed [12]. A recent study by Birck *et al* linked prophylaxis prescription data, and found similar GAS infection rates among family contacts of primary iGAS cases who did and did not receive antibiotic prophylaxis [9]. According to a recent systematic review, there is one previous study reporting GAS risk reduction after antibiotic prophylaxis, but this was during an outbreak among persons experiencing homelessness [14]. Since there is currently no general consensus as how to best prevent secondary iGAS infections in the general population, we believe our study contributes valuable insights on the benefits of antibiotic prophylaxis to prevent iGAS in high-exposure settings.

Adebanjo *et al* in 2020 summarized the available evidence on iGAS incidence among household members of primary iGAS cases [2]. The two largest previous studies found 1,240 and 4,520 iGAS cases per 100,000 person-years in the 30-day risk period among close contacts [2, 3]. Our incidence estimates for household contacts in this risk period, of 2,995 before and 575 per 100,000 person-years after the policy change, are in line with this order of magnitude.

After the COVID-19 pandemic, many countries reported an increase in iGAS incidence in 2022-2023, including the Netherlands [4]. Here, we report a population iGAS incidence of 7.07 per 100,000 person-years during 2022-2024, peaking in early 2023 at 14.15, which is roughly in line with published population incidences covering this period [10, 11]. A Danish study reported an incidence of 6.6 per 100,000 in children 0-17 years of age in 2022-2023, with the majority under the age of 5, consistent with the incidence in children in our study (Table 1) [12].

Surprisingly, we did not observe an association between iGAS risk and low household SES. Concordantly, 30-day mortality did not differ significantly by SES. Deprivation has been repeatedly shown to be an important risk factor for iGAS [1, 10, 13, 14]. Likely, our categorization of SES is not sensitive enough to capture the vulnerable populations most at risk for iGAS, such as people who inject drugs and people experiencing homelessness.

## Limitations

Our study has limitations, mainly affecting the estimated impact of the policy change on secondary iGAS incidence. The patient data submitted with the iGAS isolate did not allow us to distinguish between iGAS disease presentations for which prophylaxis was already indicated in 2022 (i.e. STSS and necrotizing fasciitis) versus newly notifiable iGAS. Especially for clusters with isolates cultured just around the date of the policy change, it is not clear if the contacts were already offered prophylaxis. One cluster had an index date before the policy change but secondary iGAS infection after. We decided to categorize this as a cluster under the old policy. Due to the low number of secondary cases, this decision impacted our results, increasing the SAR under the old policy. Also, the proportion of secondary cases occurring too soon after the primary case to receive prophylaxis may have differed between the two periods, which impacts the preventable fraction of secondary iGAS infections. The low number of (secondary) cases limited the power of our study, and precluded analyses among other types of close contacts such as non-household family members. Further, as dominant *emm* types also differed between the periods, it is not possible to fully disentangle effects of the policy change and differences in transmissibility or virulence of circulating *emm* types, despite adjusting estimates for calendar time. Lastly, the fact that 11.5% of submitted isolates could not be deterministically linked to a unique person in the population registry presents an important limitation, resulting in a lower incidence estimate. As linkage was based on postal code, birth date and sex, it is possible that twins and persons in large institutional households are overrepresented in the non-linked isolates. This might have resulted in an underestimation of the number of secondary household cases for both periods. While the median and IQR of household contacts per case were identical between the periods, the averages are very different, indicating a skewedness due to large institutional households included in the period with the new policy. Despite the adjustment for age group, this may have resulted in a more fragile population at risk included in the new policy period. On the other hand, the closeness of the contacts is likely to be less in such institutional households. The data did not allow us to distinguish between wards or rooms within institutional households, and the intensity of contact between household members may vary significantly.

The number of secondary iGAS cases was insufficient for a thorough analysis of host risk factors among the close contacts. Some countries, such as the United Kingdom, base prophylaxis eligibility not only on being a close contact to an index case but also on factors such as age, pregnancy, and viral infections [1]. In the Netherlands, the deliberate choice was made not to make such a distinction, for two reasons: to prevent ongoing household transmission through a third (non-risk group) household member, and to enhance feasibility of timely prophylaxis by removing the need to assess risk factors.

## Conclusions

In this nationwide cohort study, we observed a substantial reduction in the risk of secondary iGAS infection among household contacts after the Dutch iGAS antibiotic prophylaxis policy change. However, the overwhelming majority of iGAS occurs as sporadic cases. Therefore, while antibiotic prophylaxis likely prevents a significant disease burden in contacts at high risk, the impact on overall iGAS incidence can only be very modest.

## Data Availability

This study was performed using non-public microdata from Statistics Netherlands and laboratory data from the Netherlands Reference Laboratory for Bacterial Meningitis (Amsterdam UMC, Amsterdam, Netherlands). These data cannot be shared by the authors.

## Access to data

BdG and AvR had full access to all the data in the study and take responsibility for the integrity of the data and the accuracy of the data analysis.

## Data sharing statement

This study was performed using non-public microdata from Statistics Netherlands. These data cannot be shared by the authors.

