## Supplementary material for "The risk of household secondary invasive Group A Streptococcal infections after a prophylaxis policy change in the Netherlands": Suppl

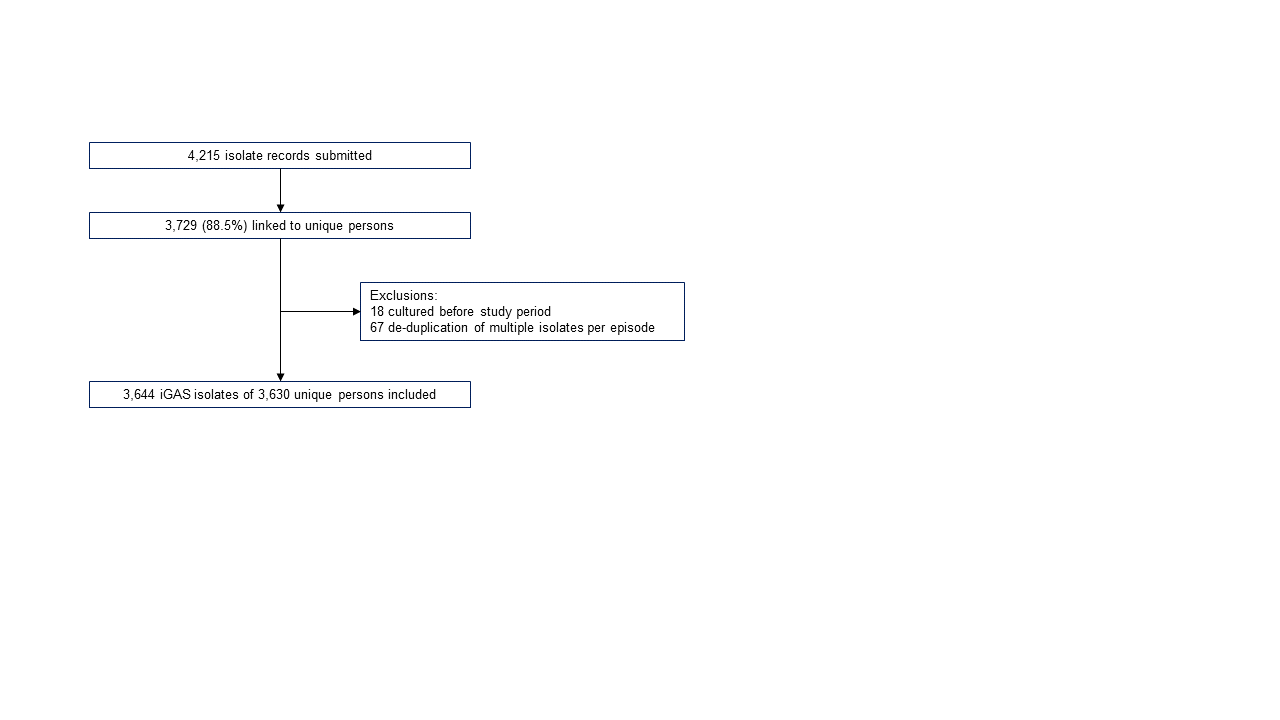

Supplementary Figure 1. Flowchart of inclusions of iGAS isolates, the Netherlands, for the study period 1 April 2022-31 December 2024.

Supplementary Table 1. Linkage of isolate data to population registry by age group, gender, policy period and *emm* type, 1 April 2022-31 December 2024, the Netherlands.

| Variable | Level | Not linked | Linked | Percentage linked | OR (95% confidence interval) |
| --- | --- | --- | --- | --- | --- |
| Age group | 0-5 | 52 | 375 | 87.8 | [ref] |
|  | 6-19 | 26 | 243 | 90.3 | 1.44 (0.86-2.41) |
|  | 20-45 | 152 | 1127 | 88.1 | 1.05 (0.73-1.47) |
|  | 46-65 | 61 | 807 | 93.0 | 2.08 (1.39-3.1) |
|  | 66+ | 65 | 1160 | 94.7 | 2.77 (1.87-4.09) |
| Sex | Male | 182 | 1713 | 90.4 | [ref] |
|  | Female or unknown | 174 | 1999 | 92.0 | 1.33 (1.06-1.67) |
| Policy | New (20 January 2023- 31 December 2024) | 249 | 2606 | 91.3 | [ref] |
|  | Old (1 April 2022- 19 January 2023) | 107 | 1106 | 91.2 | 1.04 (0.81-1.35) |
| *emm* type | 1.0 Cluster A-C3 | 103 | 1051 | 91.1 | [ref] |
|  | 1.134 Cluster A-C3 | 6 | 102 | 94.4 | 1.6 (0.74-4.18) |
|  | 12.0 Cluster A-C4 | 20 | 201 | 91.0 | 1.06 (0.65-1.81) |
|  | 22.0 Cluster E4 | 23 | 271 | 92.2 | 1.27 (0.80-2.09) |
|  | 3.93 Cluster A-C5 | 36 | 538 | 93.7 | 1.42 (0.95-2.16) |
|  | 4.0 Cluster E1 | 22 | 371 | 94.4 | 1.90 (1.20-3.14) |
|  | 87.0 Cluster E3 | 13 | 174 | 93.0 | 1.36 (0.77-2.61) |
|  | 89.0 Cluster E4 | 22 | 192 | 89.7 | 0.86 (0.54-1.44) |
|  | Other *emm* type | 111 | 812 | 88.0 | 0.71 (0.53-0.95) |

Supplementary Table 2. Characteristics of the study population.

|  |  | N (%) |
| --- | --- | --- |
| Total |  | 19,006,247 |
| Age group (years) at the start of follow-up | 0-5 | 1,559,629 ( 8.2) |
|  | 6-19 | 2,906,149 (15.3) |
|  | 20-45 | 6,308,794 (33.2) |
|  | 46-65 | 4,888,424 (25.7) |
|  | 66+ | 3,343,251 (17.6) |
| Sex | Female or unknown | 9,538,996 (50.2) |
|  | Male | 9,467,251 (49.8) |
| Household socioeconomic quintile | 1 | 3,697,898 (20.0) |
|  | 2 | 3,697,894 (20.0) |
|  | 3 | 3,697,536 (20.0) |
|  | 4 | 3,697,587 (20.0) |
|  | 5 | 3,697,546 (20.0) |

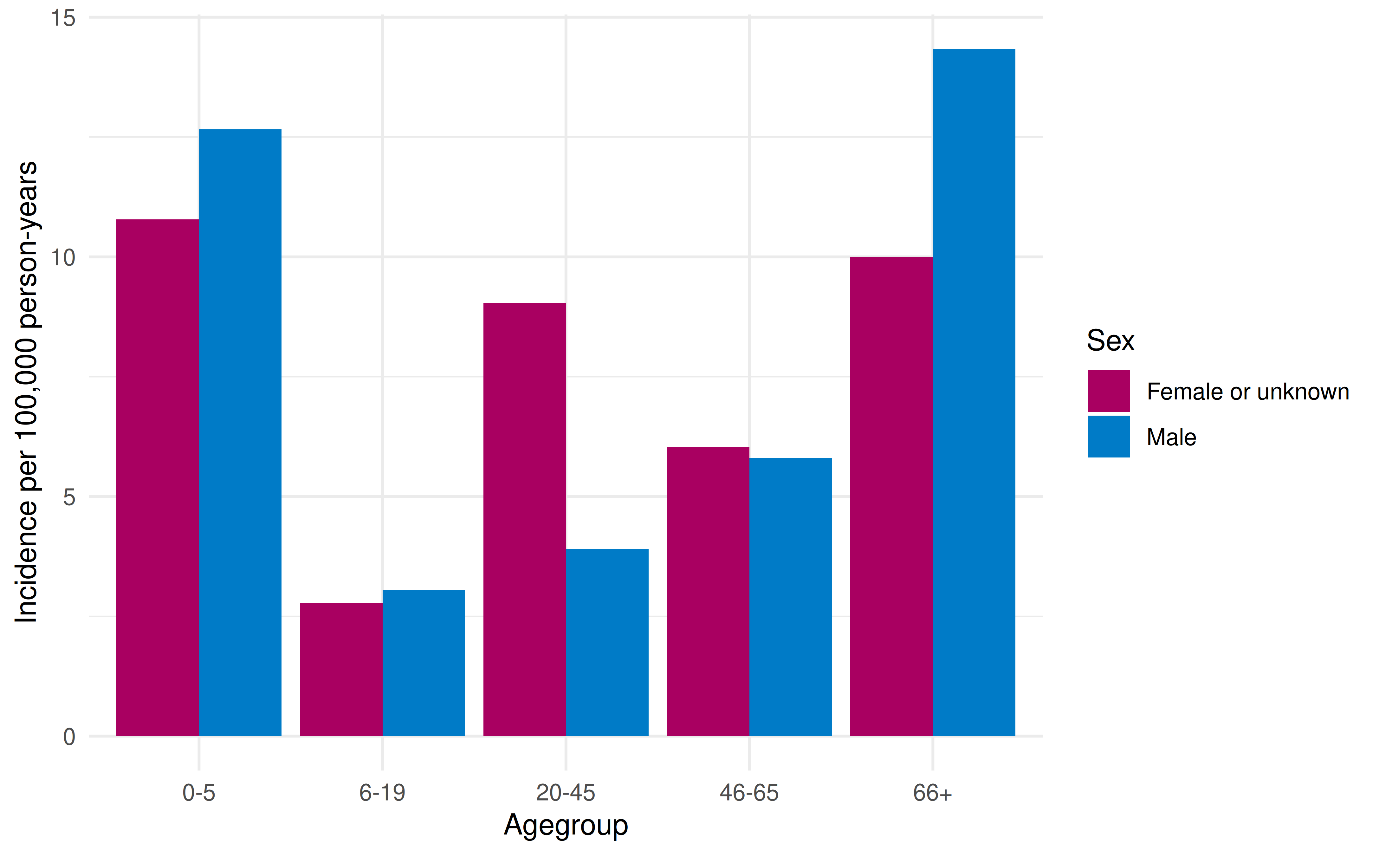
Supplementary Figure 2. Population iGAS incidence per 100,000 by age group and sex, 1 April 2022- 31 December 2024, the Netherlands.
